# Protection by 4th dose of BNT162b2 against Omicron in Israel

**DOI:** 10.1101/2022.02.01.22270232

**Authors:** Yinon M. Bar-On, Yair Goldberg, Micha Mandel, Omri Bodenheimer, Ofra Amir, Laurence Freedman, Sharon Alroy-Preis, Nachman Ash, Amit Huppert, Ron Milo

**Affiliations:** Department of Plant and Environmental Sciences, Weizmann Institute of Science, Israel; Technion - Israel Institute of Technology, Israel; The Hebrew University of Jerusalem, Israel; Israel Ministry of Health, Israel; The Bio-statistical and Bio-mathematical Unit, The Gertner Institute for Epidemiology & Health Policy Research, Sheba Medical Center, Israel; The Faculty of Medicine, Tel Aviv University, Israel

## Abstract

**BACKGROUND:** On January 2, 2022, Israel began administering a fourth dose of BNT162b2 vaccine (Pfizer-BioNTech) to people aged over 60 years and at-risk populations, who had received a third dose of vaccine at least 4 months earlier. The effect of the fourth dose on confirmed coronavirus 2019 disease (Covid-19) and severe illness are still unclear.

**METHODS:** We extracted data for the Omicron-dominated period January 15 through January 27, 2022, from the Israeli Ministry of Health database regarding 1,138,681 persons aged over 60 years and eligible for the fourth dose. We compared the rate of confirmed Covid-19 and severe illness between those who had received a fourth dose at least 12 days earlier, those who had received only three doses, and those 3 to 7 days after receiving the fourth dose. We used Poisson regression after adjusting for possible confounding factors.

**RESULTS:** The rate of confirmed infection was lower in people 12 or more days after their fourth dose than among those who received only three doses and those 3 to 7 days after vaccination by factors of 2.0 (95% confidence interval [CI], 2.0 to 2.1) and 1.9 (95% CI, 1.8 to 2.0), respectively. The rate of severe illness was lower by factors of 4.3 (95% CI, 2.4 to 7.6) and 4.0 (95% CI, 2.2 to 7.5).

**CONCLUSIONS:** Rates of confirmed Covid-19 and severe illness were lower following a fourth dose compared to only three doses.

## Introduction

During late December 2021, with the emergence of the Omicron variant, prevalence of both confirmed infections and severe illness rose sharply in Israel. Contributing factors were the increased immune evasion by the variant,^1^ and the passage of more than four months since most adults had received their third vaccine dose. In an effort to address the challenges presented by the Omicron variant and to reduce the load on the healthcare system, Israeli authorities approved on January 2, 2022 the administration of a fourth dose of the BNT162b2 vaccine to persons who were 60 years of age or older, as well as to high-risk populations and health-care workers. The real-world effectiveness of the fourth dose in decreasing the rate of confirmed infection and severe illness remains unclear. Here, we use data from the Israeli Ministry of Health national database to study the effectiveness of the fourth dose against confirmed infection and severe illness.

## Methods

### STUDY POPULATION

We included in the analysis individuals who, on January 1, 2022, were 60 years of age or older and had received three doses of BNT162b2 at least 4 months before the start of the study period. We excluded the following persons from the analysis: those who had died before the beginning of the study period; those with no information regarding their age or sex; those who had a confirmed SARS-CoV-2 infection using either Polymerase-Chain-Reaction (PCR) assay or a state-regulated rapid antigen test before the beginning of the study; those who had received a third dose before their age group became eligible (i.e., before July 30 2021); those who had been abroad during the entire study period (persons were considered as ‘being abroad’ during the period from 10 days before traveling to 10 days after their return to Israel); and those who received a vaccine dose from a brand other than Pfizer-BioNTech. A total of 1,138,681 participants met these inclusion criteria (Figure 1).

**Figure 1.**
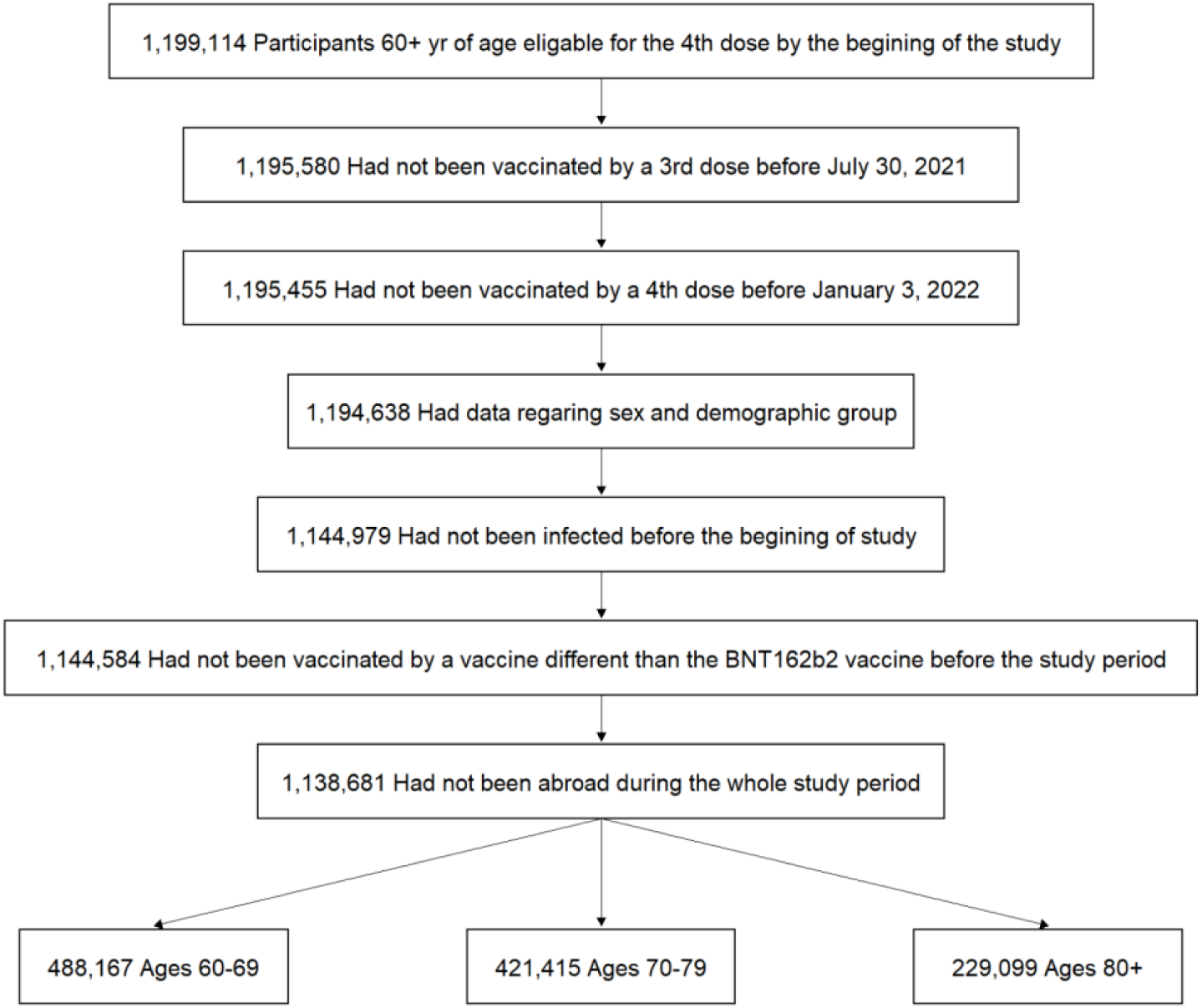
Study population. The participants in the study included persons who were 60 years of age or older, who were not infected by SARS-CoV-2 before the study period, and were eligible for the 4th dose at the beginning of the study, had available data regarding sex and demographic sector, had not stayed abroad during the whole study period, and had not been vaccinated with a vaccine different fromBNT162b2 before the study period. Age groups as of January 1, 2022.

For those who met the inclusion criteria, we extracted information covering the study period January 15 through January 27, 2022 regarding SARS-CoV-2 infection (confirmed either by state-regulated rapid antigen tests or by PCR) and severe illness due to Covid-19 (defined using the NIH definition^2^ as a resting respiratory rate of more than 30 breaths per minute, an oxygen saturation of less than 94% while breathing ambient air, or a ratio of partial pressure of arterial oxygen to fraction of inspired oxygen of less than 300). During this period, infections were overwhelmingly dominated by the Omicron variant. We also extracted data regarding vaccination (dates and brands of first, second, third and fourth doses), demographic variables such as age, sex, and demographic group (general Jewish, Arab, or ultra-Orthodox Jewish population) as determined by the individual’s statistical area of residence (similar to a census block^3^).

### STUDY DESIGN

The study period started on January 15, 2022 and ended on January 27, 2022 for confirmed infection and on January 21, 2022 for severe illness. The starting date was set to 12 days after the start of the vaccination campaign (January 3, 2022), so that all study groups would be represented throughout the study period (Figure S1 in Supplementary Appendix). The end dates were designed to minimize the effects of missing outcome data due to delays in reporting test results and due to time from diagnosis to the development of severe illness.

The design was similar to a previous study of the protection conferred by the third vaccine dose compared to the second dose.^4^ We calculated the total number of person-days at risk, and the incidence of confirmed infection and of severe illness due to Covid-19, in one treatment group and two control groups. The treatment group was composed of participants for whom 12 days or more elapsed from receipt of the fourth dose. The first control group included individuals who were eligible for a fourth dose but had not yet received it. As this natural control group might differ from the treatment group by unmeasured confounding variables, a second control group was defined as individuals for whom 3-7 days had passed since receiving the fourth dose. This control group included the same individuals as the treatment group, but at times when the fourth dose was not expected to be effective. The time of onset of severe Covid-19 was assigned to be the date of the test confirming infection. Data was retrieved on January 29, 2022, eight days after the end of the study period, allowing at least 7 full days of follow-up time for the development of severe illness. To allow for the same followup for all individuals, we considered participants to have severe illness if they were hospitalized in severe condition within seven days of their confirmed infection.

### OVERSIGHT

The study was approved by the institutional review board of the Sheba Medical Center. All the authors contributed to conceiving the study, critically reviewed the results, approved the final version, and made the decision to submit the manuscript for publication. The Israeli Ministry of Health and Pfizer have a data-sharing agreement, but only the final results of this study were shared.

### STATISTICAL ANALYSIS

Using Poisson regression, we calculated the rates of confirmed infection and severe illness due to Covid-19 per 100,000 person-days at risk for each dynamic cohort (including the three study groups as factors in the model), adjusting for the following demographic variables: age group (60-69, 70-79, and 80+ years), sex, and demographic group (general Jewish, Arab, ultra-Orthodox Jewish). Since incidences of both confirmed infection and severe illness increased rapidly during January 2022, days at the beginning of the study period had lower exposure risk compared to days at the end. Moreover, the fraction of the population in each dynamic cohort also changed throughout the study period (Figure S1 in the Supplementary Appendix). Therefore, we included calendar date as an additional covariate to account for growing exposure risk.^5^

For each outcome, we estimated the incidence rate in three groups of participants: eligible individuals who had not received the fourth dose, individuals for whom 3-7 days had passed since receiving the fourth dose, and individuals for whom 12 days or more had passed since receiving the fourth dose. We calculated two incidence rate ratios for each outcome. First, we compared the group of eligible individuals who did not receive the fourth dose with the group of individuals for whom 12 days or more had passed since receiving the fourth dose. Second, we compared the group of individuals for whom 3-7 days had passed since receiving the fourth dose with the group of individuals for whom 12 days or more had passed since receiving the fourth dose. In addition, adjusted rate differences per 100,000 person days during the study period were calculated in a way similar to that used by Bar-On et al.^5^ Confidence intervals were calculated by exponenting the 95% confidence intervals for the regression coefficients without adjustment for multiplicity.

To further examine the change in the rate of confirmed infection as a function of time since receipt of the fourth dose, we fitted a Poisson regression with days after the fourth dose as factors in the model. The period before receipt of the fourth dose was used as the reference category. This analysis was similar to our previous analysis of the effectiveness of the third dose,^5^ and produced rate ratios for different days after the fourth vaccine dose.

To account for possible biases, we performed several sensitivity analyses. These are described in the Supplementary Appendix.

## Results

### STUDY POPULATION

The total number of events and person-days at risk for each of the 3 comparative groups, along with the distribution of covariates used in the analysis are presented in Table 1. Overall, the distributions of covariates for persons 12 days or more after receiving the fourth dose are similar to those 3-7 days from vaccination. Compared with those who had not yet received a fourth dose, the groups of those that received a fourth dose (3-7 days post-vaccination and 12+ days post vaccination) included more person-days over the age of 80 (24.2% and 26.9% vs. 16.3%) and more person-days from the general Jewish population (95.0% and 92.8% vs. 85.6%). Those who had not yet received a fourth dose had almost twice the number of risk days compared to persons 12 days or of more after receiving the fourth dose (7.6 millions compared to 3.4 millions), and had many more confirmed infections (42,693 vs. 9071) and severe cases (195 vs. 13).

**Table 1:** Demographic and clinical characteristics of the different cohorts. The table presents the proportion of person-days at risk (for the confirmed infection analysis) instead of the number of individuals, as people can move between cohorts. Risk days and infections are calculated for the study period January 15-27, 2022. Severe disease cases are limited to a 7-day followup duration for infections that occurred during the period January 15-21, 2022.

| Group | 3rd dose<br>Person-days at risk = 7,603,132 |  |  | 4th dose 3-7 days<br>Person-days at risk = 1,264,767 |  |  | 4th dose 12+ days<br>Person-days at risk = 3,421,826 |  |  |
| --- | --- | --- | --- | --- | --- | --- | --- | --- | --- |
|  | % person days at risk | # infections | # severe Covid-19 | % person days at risk | # infections | # severe Covid-19 | % person days at risk | # infections | # severe Covid-19 |
| Female | 54.4% | 23,160 | 85 | 54.0% | 3,049 | 29 | 50.6% | 4,222 | 6 |
| Male | 45.6% | 19,533 | 110 | 46.0% | 2,896 | 26 | 49.4% | 4,849 | 7 |
| 60-69 | 51.1% | 26,329 | 30 | 32.9% | 2,356 | 3 | 32.1% | 3,485 | 3 |
| 70-79 | 32.6% | 11,943 | 52 | 40.2% | 2,154 | 15 | 43.7% | 3,484 | 5 |
| 80+ | 16.3% | 4,421 | 113 | 26.9% | 1,435 | 37 | 24.2% | 2,102 | 5 |
| General Jewish | 85.6% | 36,371 | 147 | 92.8% | 5,475 | 48 | 95.0% | 8,536 | 12 |
| Ultra-Orthodox | 4.4% | 2,157 | 15 | 2.8% | 243 | 3 | 2.6% | 283 | 0 |
| Arab | 9.9% | 4,165 | 33 | 4.4% | 227 | 4 | 2.4% | 252 | 1 |

### PROTECTION CONFERRED BY THE FOURTH DOSE

The results regarding confirmed infection and severe illness are summarized in Table 2. The rate of confirmed infection for the group of people who received the fourth dose 12 days or more previously was lower by a factor of 2.0 (95% confidence interval [CI], 2.0 to 2.1) compared to the group of eligible people who had not received the fourth dose, and was lower by a factor of 1.9 (95% CI, 1.8 to 1.9) compared to those who had received the fourth dose 3-7 days previously. The adjusted rate differences were 279 (95% CI, 271 to 287) and 234 (95% CI, 219 to 247) cases per 100,000 person-days at risk between the treatment group and the two control groups.

**Table 2.** Results of the Poisson regression analysis for confirmed infection and severe illness between the different study groups.

|  | Cases (person-days at risk) |  |  | Rate Ratio (95% CI) |  | Adjusted rate difference per 100,000 person-days at risk (95% CI) |  |
| --- | --- | --- | --- | --- | --- | --- | --- |
|  | 3rd dose only | 3-7 days after 4th dose | 12+ days after 4th dose | 3rd dose only vs. 12+ days after 4th dose | 3-7 vs. 12+ days after 4th dose | 3rd dose only vs. 12+ after 4th dose | 3-7 vs. 12+ days after 4th dose |
| Confirmed Infections | 42,693<br>(7,603,132) | 5,945<br>(1,264,767) | 9,071<br>(3,421,826) | 2.0<br>[2.0, 2.1] | 1.9<br>[1.8, 2.0] | 279<br>[271, 287] | 234<br>[219, 247] |
| Severe illness | 195<br>(4,277,639) | 55<br>(1,023,355) | 13<br>(980,984) | 4.3<br>[2.4, 7.6] | 4.0<br>[2.2, 7.5] | 3.8<br>[2.8, 4.8] | 3.5<br>[2.1, 5.1] |

The rate of severe illness for the group of people who received the fourth dose 12 days or more previously was lower by a factor of 4.3 (95% CI 2.4 to 7.6) compared to the group of eligible people who had received only three doses, and was lower by a factor of 4.0 (95% CI 2.2 to 7.5) compared to those who had the fourth dose 3-7 days previously. The adjusted rate differences were 3.8 (95% CI, 2.8 to 4.8) and 3.5 (95% CI, 2.1 to 5.1) cases per 100,000 person-days at risk compared to the two control groups, respectively.

Figure 2 shows the rate ratio of confirmed infections for those who had not received a fourth dose compared to those receiving a fourth dose on different days following that dose. During days 3-7 after vaccination, the rate ratio is near 1; it starts increasing after about one week, and reaches 2-3-fold two weeks after vaccination.

**Figure 2.**
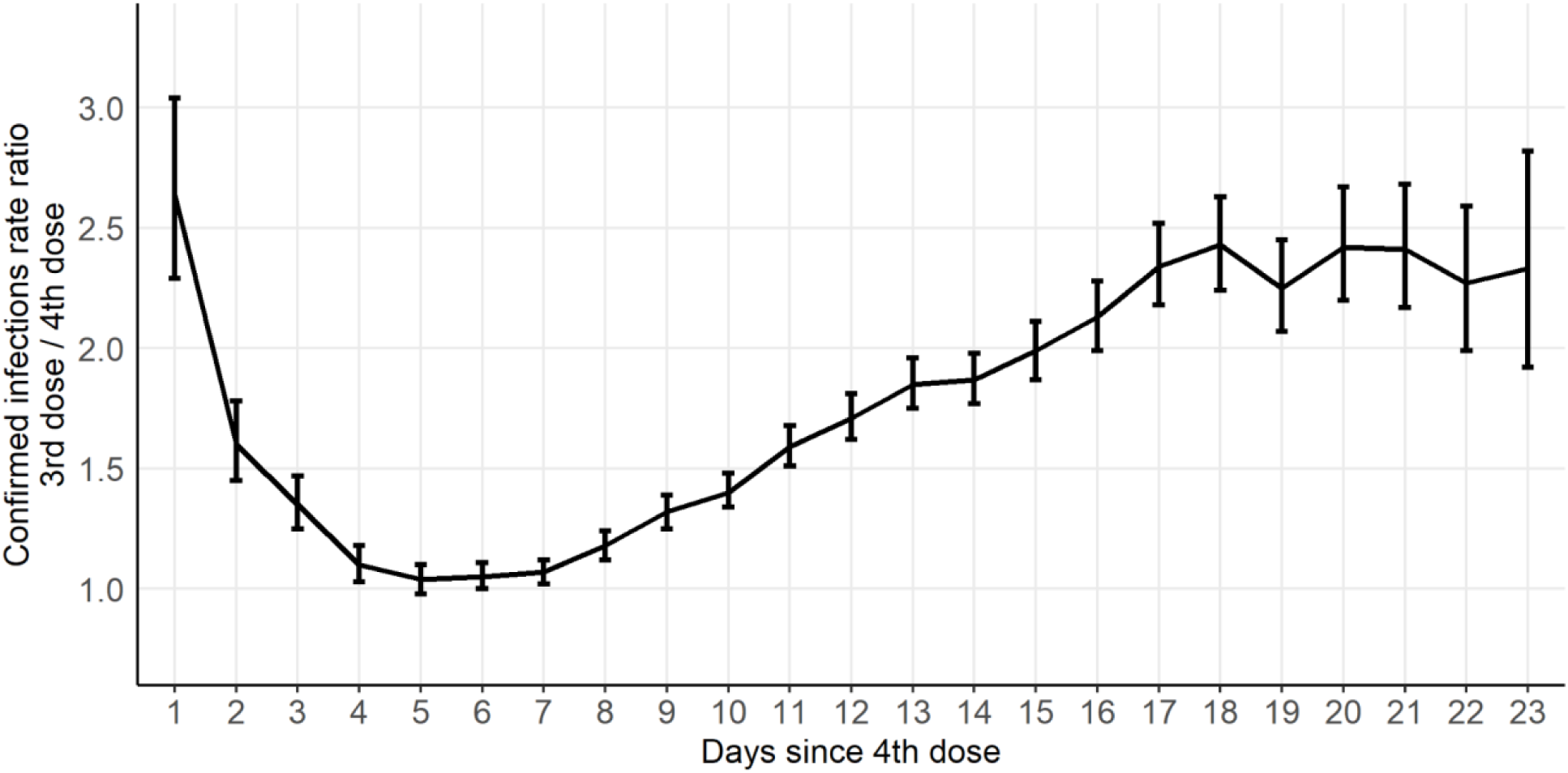
The rate ratio for confirmed infections between the group of people eligible for a fourth dose who had not yet received it to those who had received a fourth dose, as a function of time since the fourth dose.

The results of several sensitivity analyses are shown in Table S1 of the Supplementary Appendix. All analyses show the same rate ratios of about 2 and 4 for confirmed infection and severe illness, respectively.

## Discussion

The Omicron variant is genetically divergent from the ancestral SARS-CoV-2 strain for which the BNT162b2 vaccine was tailored. Nevertheless, the results presented here indicate that a fourth dose is still able to provide added protection against confirmed infection and severe illness with Omicron compared to three vaccine doses that were given at least four months previously. The incidence rate for confirmed infection was lower by a factor of two, and the rate of severe disease by a factor of four in people who received the fourth dose 12 days or more previously compared to eligible individuals who did not receive the fourth dose. While our main analysis relates only to people aged 60 years and older, an additional analysis performed for select groups of people aged 20-59 showed similar results for protection against confirmed infections (see Table S1 in the Supplementary Appendix).

Although our analysis attempts to address biases such as confounding, some sources of bias may not have been measured or adequately controlled, for example behavioral differences between people who received the fourth dose and those who did not. For severe illness, differences in the prevalence of comorbidities could potentially affect the results, but are not recorded in the national database. To address some of these biases, we compared the rate of confirmed infection and severe illness within the group of people who received the fourth dose. Specifically, the rate of confirmed infections and severe illness on days 3-7 following vaccination, before the vaccine is expected to take effect,^5^ was compared to the rate 12 days or more after vaccination. The rate ratios obtained were very similar to those obtained when comparing the latter group to people who did not receive the fourth dose. Thus, the results appear to be robust to the selection of people who opted to receive the fourth dose. Figure 2 that compares the rate ratio over time suggests that the protection of the fourth dose continues to increasee beyond day 12, reaching a rate ratio of about 2.5 after 2.5-3 weeks. Longer follow-up will help to evaluate the protection of the fourth dose over longer periods of time.

In addition, several sensitivity analyses were performed to assess the robustness of the results to further potential biases. First, we performed the analyses using data only from the general Jewish population, since the participants in that group are enriched in the population that received the fourth dose. Second, we included in the model the exposure at the place of residence. The results of these analyses, presented in Table S1 of the Supplementary Appendix, are similar to the results of the main analysis. As discussed in the Supplementary Appendix, the testing policy in Israel was changed before the study period in early January. Under the new guidelines, vaccinated people under the age of 60 who were exposed to a confirmed case or developed symptoms were asked to perform a rapid antigen test at home, and if positive to perform a state-regulated antigen test. The guidelines for the elderly population have not been changed, and they were asked to perform PCR tests, but the change in the under-60s could indirectly affect the testing behavior in the 60+ age group. To test the possible effect of the type of diagnostic test, we repeated the analysis using only positive PCR tests to confirm infections. As shown in Table S1 in the Supplementary Appendix, using only PCR tests resulted in very minor changes to the estimated level of protection conferred by the fourth dose. In addition, we compared the testing rate and test type (PCR or antigen) of people who received the fourth dose compared to those who received only three doses, and found the differences to be of limited extent and such that a bias, if existed, could be towards underestimation of the level of protection (Figure S2 in the Supplementary Appendix).

Overall, these analyses provide evidence for the effectiveness of a fourth vaccine dose against both confirmed infection and severe illness with the Omicron variant, compared to a third dose that was administered more than four months previously. Several reports have indicated that the protection from hospital admission by a third dose that was given more than three months previously is substantially lower with the Omicron variant compared to the protection of a fresh third dose against the Delta variant.^1,6,7^ The results of this study suggest that a fourth dose could increase protection against severe illness relative to three doses that have been administered over four months ago. Giving the fourth dose to individuals who were at risk to develop severe disease has been instrumental in limiting the burden on hospitals in Israel during the fast and wide-spreading Omicron surge.

## Data Availability

The individual-level data used in this study cannot be publicly shared even if anonymized due to privacy restrictions.

## Supplementary Appendix

### Supplementary Methods - Database

The Ministry of Health (MOH) in Israel collects all COVID-19 related variables in a central database. These include data on all PCR and antigen tests and results, vaccination dates and type (almost all received the Pfizer-BioNTech vaccine), daily clinical status of all COVID-19 hospitalized patients, and COVID-19 related deaths. Specifically, the data used for conducting this study included vaccination dates (second and third doses), PCR tests (dates and results), hospital admission dates (if relevant), clinical severity status (severe illness or death), and demographic variables such as age, sex, and demographic group (General Jewish, Arab, ultra-Orthodox Jewish). Severe disease is defined as a resting respiratory rate >30 breaths per minute, oxygen saturation on room air <94%, or ratio of PaO2 to FiO2 <300. Those who died from COVID-19 during the follow-up period were also counted as severe disease cases in our analysis. The fact that Israel has a central health care system increases the coverage and reliability of the data. A small fraction of the population with missing observations on gender or demographic sector were excluded from our analysis. They comprised ≈0.1% of the total population and were most likely missing those variables at random. We also excluded from the analysis individuals for whom the area of residence was unknown, which amounts to about 2.5% of the total population in the database. As we show in Supplementary Analysis 2, this exclusion has a negligible effect on the results of the analysis. The MOH database comprises data from multiple sources. These include all MOH-approved laboratories performing PCR and antigen testing in Israel, including private laboratories, hospitals and the four Health Maintenance Organizations (HMOs) that together insure the entire Israeli population. Quality assurance of data was performed extensively over the course of the pandemic, and the data are monitored daily by the MOH, and are continuously used for public health decision-making.

PCR and antigen testing for SARS-CoV-2 is free-of-charge and widely available in Israel. Testing is required for symptomatic persons (e.g., with fever or acute respiratory illness), people who were in close contact with an infected individual, or travelers returning from abroad. When undergoing a test, persons are required to provide their unique identification number. A nasal or nasopharyngeal swab is collected and sent to a certified laboratory where it is tested (using national testing standards) either by reverse transcription quantitative PCR or using a rapid antigen test. All sampling laboratories digitally report the data to the MOH database. Turn-around intervals between nasopharyngeal sampling and test result are 48 hours at most and typically within 24 hours. Surveillance of COVID-19-associated hospitalizations is continuously performed by the MOH. Data from all hospitals are updated daily, and often twice a day. In accordance with national guidelines, healthcare providers report all hospitalizations and deaths among individuals with laboratory-confirmed SARSCoV-2 infection.

On January 7, 2022, the Israeli ministry of health changed its policy regarding the type of testing people exposed to COVID-19 cases are required to perform to exit quarantine. For vaccinated people above the age of 60, which is the population studied here, policy had not changed. Nevertheless, vaccinated people younger than 60 years of age are no longer required to perform a PCR test but can use an at-home or a state-regulated rapid antigen test. If negative, they can avoid entering quarantine. At the same time, due to the rapid increase in the incidence of COVID-19 cases throughout January 2022, PCR testing capacity in Israel became strained, which led more and more people to use state-regulated antigen tests for diagnosis. Even though the testing policy for our study group hasn’t changed dramatically, due to the large overall changes in the testing policy, we consider either a positive antigen or a positive PCR test as a confirmed infection.

While including state-regulated rapid antigen tests expands our coverage of the diagnosis tests performed by the general population, we do not cover tests performed using at-home rapid antigen kits. It is probable that with increasing incidence of COVID-19, followed by long waiting times in testing facilities, more people have opted to perform at-home rapid antigen tests. As these tests are not regulated, it is hard to discern what fraction of the people who test positive in these at-home tests report their positive result or get a PCR confirmatory test. Specifically, we do not know how similar non-reporting rates are between the study groups.

### Supplementary Analysis - Additional analyses

#### TESTING RATES BETWEEN STUDY GROUPS

One possible confounder that can affect the results of the analysis is a behavioral difference between the study groups in their tendency to perform tests for diagnosis of infection. On the one hand, it is possible that people who were more recently vaccinated feel more protected against infection and thus would be less likely to get tested in case of exposure. On the other hand, vaccination status itself may be associated with traits such as increased awareness of the pandemic, which could lead people who chose to get the fourth vaccine first to get tested more regularly. To test the possible direction and extent of such between group differences, we calculated the total number of tests performed per 100,000 people in the week between January 16 and January 23, 2022 in two groups of individuals. The first group was defined as eligible individuals who did not receive the fourth dose by January 23, 2022. The second group were individuals who received the fourth dose before January 16. The results, presented in Figure S2, indicate that the testing rate in the group of eligible people who did not receive the fourth dose was lower by about 30% than in the group of people who received a fourth dose (≈17,000 tests per 100,000 compared with ≈22,000). The fraction of rapid antigen tests out to the total tests performed by people who received the fourth dose was 13% compared to 23% in people who received only three doses. In theory, this difference could lead to an underestimate of the effect of the fourth dose in reducing the rate of confirmed infections. We use our secondary analysis, in which both our treatment group and our control group received a fourth dose, as a proxy for the effect of this difference in testing rate between recipients of the fourth dose and those that received only three doses. We obtained very similar results to our primary analysis regarding the level of protection against confirmed infection, suggesting that the effect of testing rate on the results is not substantial.

#### SENSITIVITY ANALYSES

To account for possible biases, we performed several sensitivity analyses. First, we performed the same analyses as described in the statistical analysis section using data only from PCR tests without inclusion of state-regulated antigen tests as confirmed infection. Second, the analyses were done using data only on the general Jewish population which had the highest rates of fourth dose vaccination. Third, we analyzed the data while accounting for exposure over time of each individual. This was done by binning the incidence rate per 1000 residents in each area of residence into 10 quantiles and using these quantiles as a covariate. We also fitted the model to individuals aged 20-59 who received the fourth dose (mostly health workers and people with preexisting health conditions). Due to the small number of severe cases in these ages, and since not all individuals in this age group were eligible to receive the fourth dose, we compared the rate of confirmed infection of vaccinees 3-7 days after to 12 days or more after vaccination. The results of all these analyses appear in Table S1.

**Figure S1.**
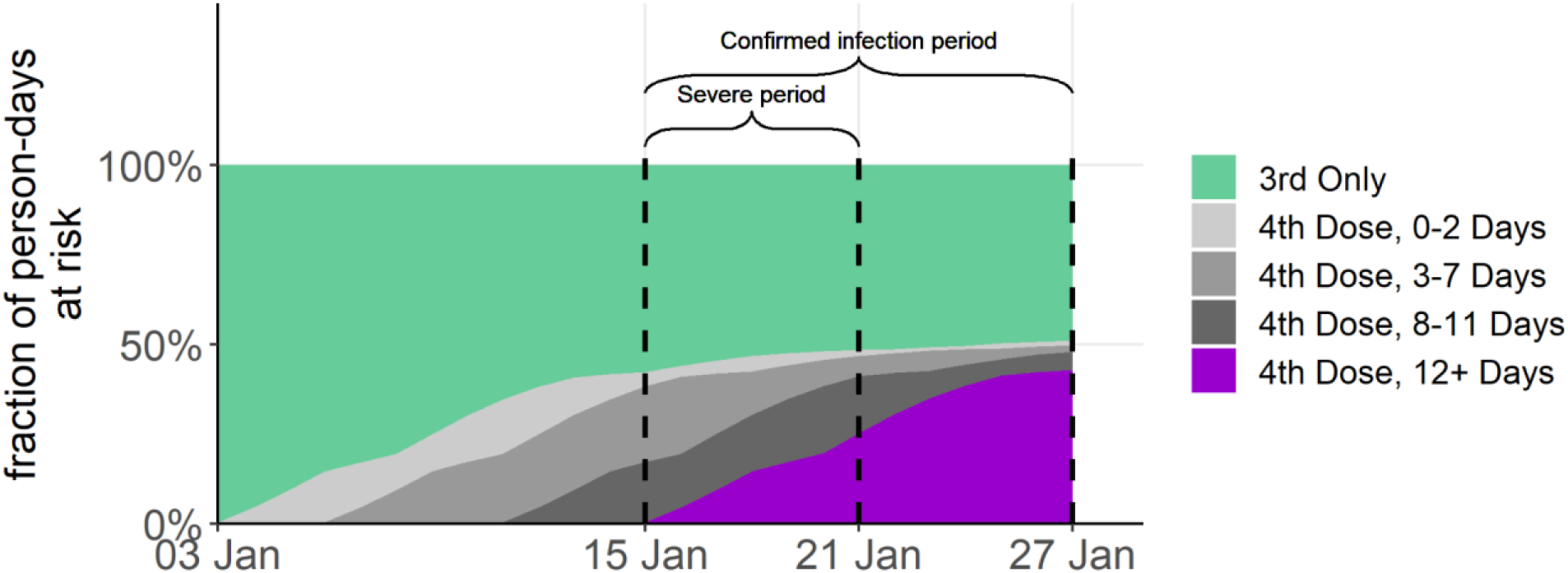
Vaccination dynamics of people aged 60 or above who were eligible for the fourth vaccine. The dashed vertical lines represent study periods for severe and confirmed infections. For both outcomes, the study period starts on January 15, 2022. For confirmed infections the study period ends on January 27, 2022. For severe illness only confirmed infections that occur before January 21, 2022 were considered.

**Figure S2.**
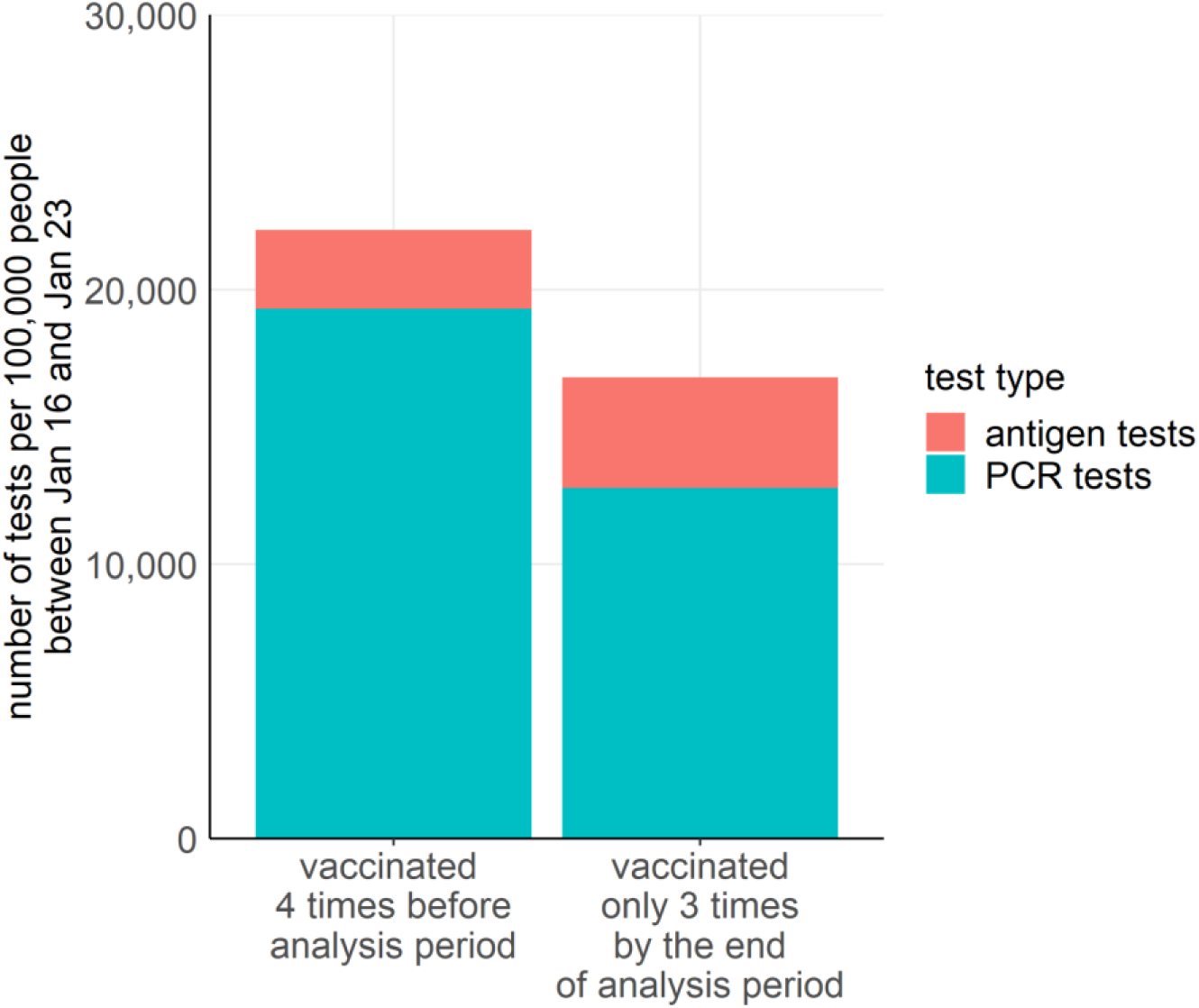
Numbers of total PCR and antigen tests per 100,000 people that were performed during January 16, 2022 and January 23, 2022 by people who received four vaccine doses before this period, and by those who, at the end of this period, were eligible for a fourth dose but had not received it.

**Table S1.**
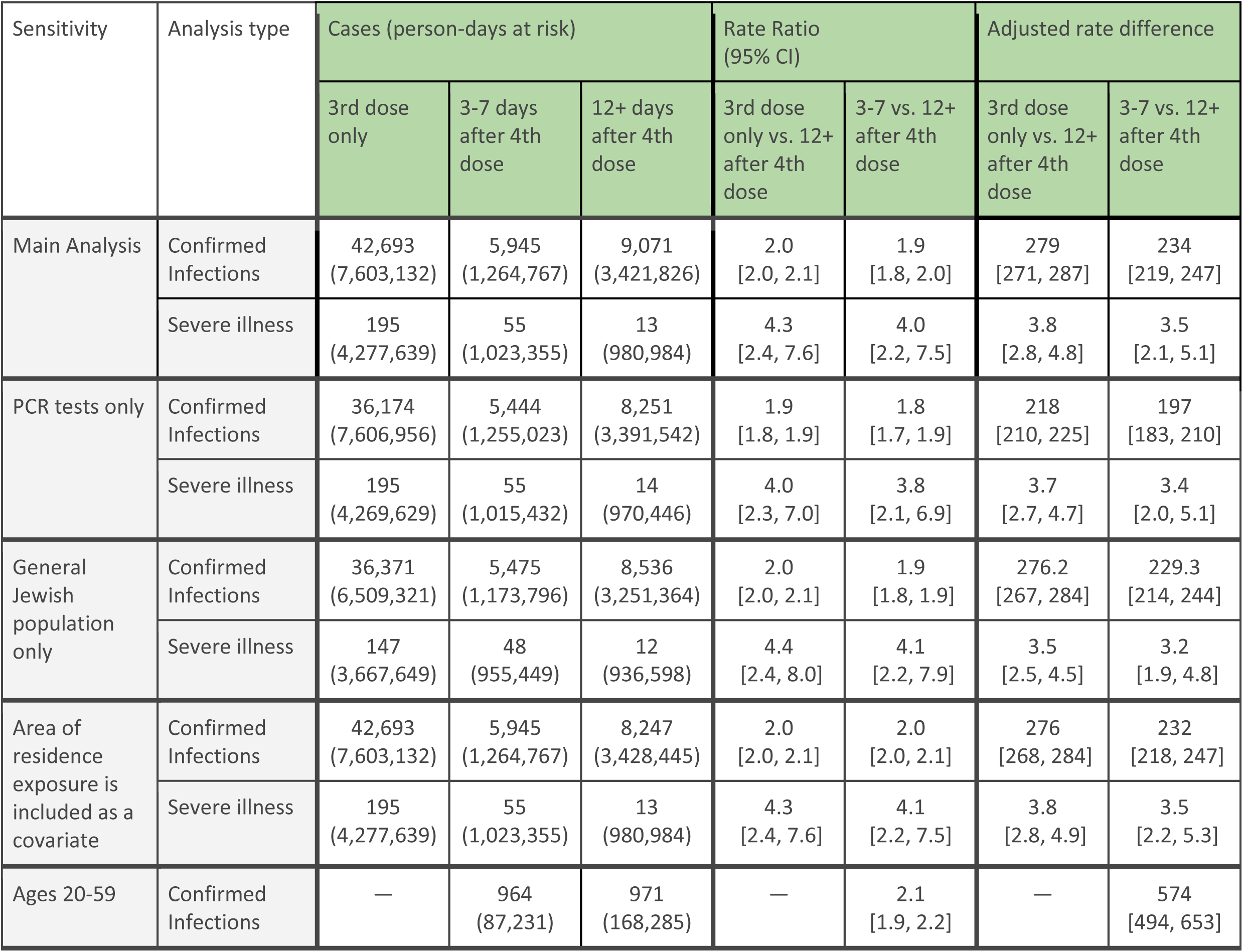
Sensitivity analyses for the main results. The first sensitivity analysis shows the results when using only positive PCR tests for confirmation of infection. The second sensitivity analysis uses only the data of the general Jewish sector. The third sensitivity analysis accounts for the exposure of each individual over time. The fourth sensitivity analysis compares the rates of confirmed infection between the early 4th dose cohort (3 to7 days after receiving the fourth dose) and the 12+ days cohort.

## References

1. SARS-CoV-2 variants of concern and variants under investigation in England - Technical briefing 19 [Internet]. Public Health England; 2021. Available from: https://assets.publishing.service.gov.uk/government/uploads/system/uploads/attachment_data/file/1005517/Technical_Briefing_19.pdf

2. Clinical Spectrum [Internet]. COVID-19 Treat. Guidel. [cited 2021 Apr 7];Available from: https://www.covid19treatmentguidelines.nih.gov/overview/clinical-spectrum/

3. Muhsen K, Na’aminh W, Lapidot Y, et al. A nationwide analysis of population group differences in the COVID-19 epidemic in Israel, February 2020–February 2021. Lancet Reg Health - Eur 2021:7:100130.

4. Bar-On YM, Goldberg Y, Mandel M, et al. Protection of BNT162b2 vaccine booster against covid-19 in israel. N Engl J Med 2021;

5. Bar-On YM, Goldberg Y, Mandel M, et al. Protection against Covid-19 by BNT162b2 Booster across Age Groups. N Engl J Med 2021:385(26):2421–30.

6. Effectiveness of a Third Dose of mRNA Vaccines Against COVID-19–Associated Emergency Department and Urgent Care Encounters and Hospitalizations Among Adults During Periods of Delta and Omicron Variant Predominance — VISION Network, 10 States, August 2021–January 2022 | MMWR [Internet]. [cited 2022 Jan 31];Available from: https://www.cdc.gov/mmwr/volumes/71/wr/mm7104e3.htm?s_cid=mm7104e3_x

7. BNT162b2 (Pfizer–Biontech) mRNA COVID-19 Vaccine Against Omicron-Related Hospital and Emergency Department Admission in a Large US Health System: A Test-Negative Design by Sara Y. Tartof, Jeff M. Slezak, Laura Puzniak, Vennis Hong, Fagen Xie, Bradley K. Ackerson, Srinivas R. Valluri, Luis Jodar, John M. McLaughlin :: SSRN [Internet]. [cited 2022 Jan 31];Available from: https://papers.ssrn.com/sol3/papers.cfm?abstract_id=4011905

